# SARS-CoV-2 genetic variations associated with COVID-19 severity

**DOI:** 10.1101/2020.05.27.20114546

**Authors:** Pakorn Aiewsakun, Patompon Wongtrakoongate, Yuttapong Thawornwattana, Suradej Hongeng, Arunee Thitithanyanont

## Abstract

Herein, we performed a genome-wide association study on SARS-CoV-2 genomes to identify genetic variations that might be associated with the COVID-19 severity. 152 full-length genomes of SARS-CoV-2 that were generated from original clinical samples and whose patient status could be determined conclusively as either “asymptomatic” or “symptomatic” were retrieved from the GISAID database. We found that nucleotide variations at the genomic position 11,083, locating in the coding region of non-structural protein 6, were associated with the COVID-19 severity. While the 11083G variant (i.e. having G at the position 11,083) was more commonly found in symptomatic patients, the 11083T variant appeared to associate more often with asymptomatic infections. We also identified three microRNAs that differentially target the two variants, namely miR-485-3p, miR-539-3p, and miR-3149. This may in part contribute to the differential association of the two SARS-CoV-2 variants with the disease severity.

## Introduction

Severe acute respiratory syndrome coronavirus 2 (SARS-CoV-2), the causative agent of coronavirus disease 2019 (COVID-19), was first reported in Wuhun, Hubei, China in late December 2019 [1,2]. SARS-CoV-2 is a positive-sense single-stranded RNA virus in the family *Coronaviridae* [3]. It is the seventh known coronavirus capable of infecting human, after HCoV-229E, HCoV-OC43, HCoV NL63, HKU1, MERS-CoV, and the original SARS-CoV. The former four typically cause non-lethal mild upper respiratory diseases, while the latter two and SARS-CoV-2 can cause severe lethal respiratory illnesses [4,5].

On 30 January 2020, approximately one month after the first reported outbreak of SARS-CoV-2 in China, the virus was found to spread to 19 countries, and the World Health Organization (WHO) declared the outbreak to be a Public Health Emergency of International Concern [6]. After the virus was found to spread to 114 countries, the WHO recognised COVID-19 as a pandemic on 11 March 2020, the first one to be caused by a coronavirus [7]. As of now (27 May 2020), the virus had spread to 213 countries and territories around the world, infecting more than 5,700,000 people [8], and this rapid surge of patients had quickly overwhelmed hospitals in many countries. Although the case-fatality ratio of SARS-CoV-2 (~3–6% [8,9]) is lower than those of SARS-CoV (11%) [10,11] and MERS-CoV (34–37%) [12], due to the great number of infected cases, the number of deaths caused by SARS-CoV-2 is much greater than those by SARS-CoV and MERS-CoV. To date, at least 350,000 deaths had been reported to be associated with SARS-CoV-2 infection [8].

The incubation period for COVID-19 is ~4–5 days, with most cases (97.5%) develop symptoms within 11–12 days of infection [13]. However, studies have reported that 5–80% of infected cases might be asymptomatic [14]. Several asymptomatic (and presymptomatic) transmissions have been reported [15], suggesting roles of asymptomatic infections in the transmission and spread of the disease. Indeed, it has been proposed that “asymptomatic carriers is a challenge to containment” [16] and that “asymptomatic transmission of SARS-CoV-2 is the Achilles’ heel of Covid-19 pandemic control” [17]. To effectively combat with the spread of the disease, this is therefore important to understand the COVID-19 pathogenesis, and its underlying factors.

Several host factors that are positively correlated with the COVID-19 severity have been identified, including patient age [9,18], low level of CD4^+^ and CD8^+^ T cell counts, and the high levels of IL-6 and IL-8 [19]. On the other hand, viral factors associated with the COVID-19 severity are still yet to be determined. There are currently more than 30,000 genomes of SARS-CoV-2 sampled from around the globe made publicly available on the database of the Global Initiative on Sharing All Influenza Data (GISAID) initiative [20], and many of these sequences had patient data. This enabled us to examine viral genetic variations that might be correlated with the severity of COVID-19 on the global scale. In this study, we performed a genome-wide association study (GWAS) on 152 SARS-CoV-2 genomes to identify potential viral genetic variations that might be associated with the COVID-19 severity.

## Results and discussion

### SARS-CoV-2 genome sequences with patient status

SARS-CoV-2 genomes with patient status were retrieved from GISAID [20]. At the time of this study, 152 genome sequences were full-length (>29,000 nucleotides (nt)), generated from original clinical samples, and had unambiguous patient status, which could be confidently determined either as “asymptomatic” or “symptomatic” (see **Materials and Methods**). To allow for accurate identification of viral genetic factors associated with COVID-19 severity, only these sequences were analysed. Together with 500 randomly sampled SARS-CoV-2 genomes from GISAID, all which were full-length and had a high sequencing coverage (but did not have clear patient information), we reconstructed a maximum likelihood (ML) phylogeny to examine how these 152 SARS-CoV-2 isolates are related to one another and to other sequences. We found that they covered a wide diversity of SARS-CoV-2, distributing across the entire tree (**Figure 1**). According to the classification scheme and method described in [21], these 152 genomes comprised 16 distinct lineages of SARS-CoV-2 (**Table 1**). The distribution of asymptomatic and symptomatic SARS-CoV-2 were different across lineages however (X^2^ test: score = 50.67, degree of freedom = 28, p-value = 0.005). While symptomatic SARS-CoV-2 were found across almost all 16 lineages, majority of asymptomatic SARS-CoV-2 in this dataset were predominantly found to be those of lineage B and B.5. Mirroring this observation, asymptomatic viruses in this dataset were mostly isolated from patients exposed to the virus in Japan (60/72 = 83.33%) and India (6/72 = 8.33%), while symptomatic cases were found around the globe (**Table 2**).

**Figure 1.**
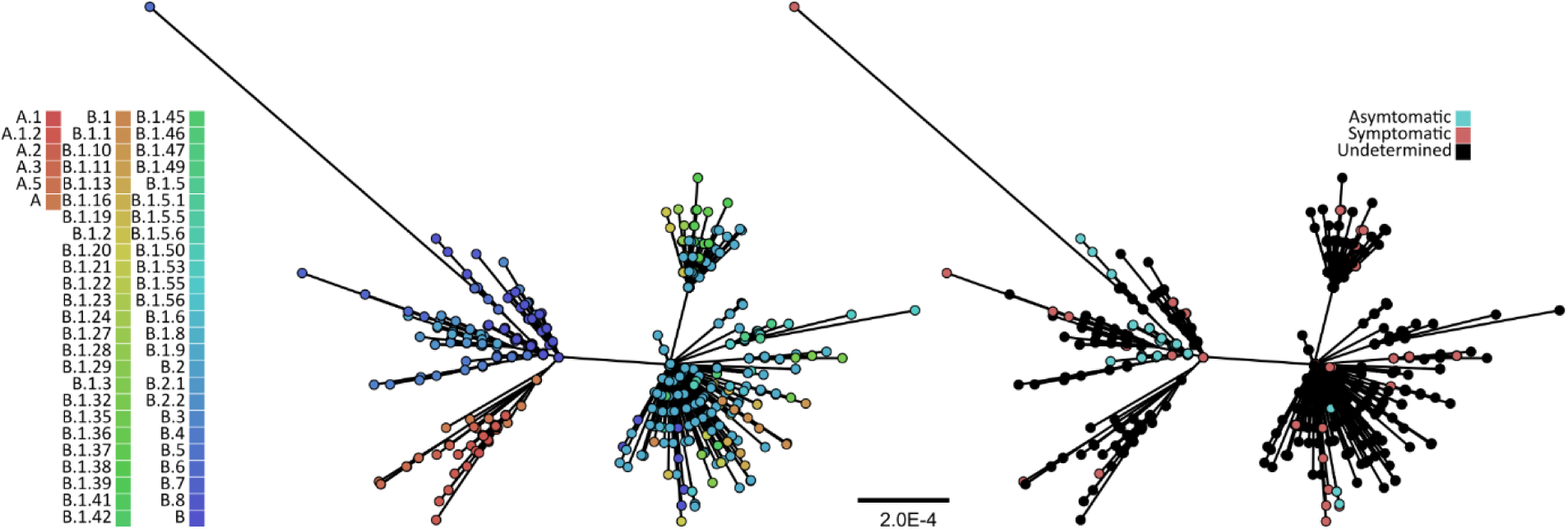
Maximum likelihood phylogeny of 625 SARS-CoV-2 full-length genomes. The tree was reconstructed using IQ-TREE [22] and the GRT^+^I nucleotide substitution model, the best-fit model as determined under the Bayesian information criterion by ModelFinder [23]. The bootstrap clade support values were computed based on 1,000 pseudoreplicate datasets, and only branches with >70% bootstrap support are shown. The scales bar is in the units of substitutions per site. The tips were coloured either by their lineages (left) as identified by pangolin (github.com/hCoV-2019/pangolin), or by COVID-19 severity (right).

**Table 1:** Lineages of the 152 SARS-CoV-2 genomes with patient status information according to the classification scheme and method described in [21].

| Severity \ Lineage | A | A.1 | A.2 | A.3 | B | B.1 | B.1.22 | B.1.3 | B.1.36 | B.1.5 | B.1.6 | B.2 | B.3 | B.5 | B.6 | B.7 | Total |
| --- | --- | --- | --- | --- | --- | --- | --- | --- | --- | --- | --- | --- | --- | --- | --- | --- | --- |
| Asymptomatic | 1 | 1 | 1 | 0 | 50 | 4 | 0 | 0 | 0 | 0 | 1 | 0 | 0 | 14 | 0 | 0 | 72 |
| Symptomatic | 0 | 4 | 0 | 1 | 17 | 37 | 1 | 2 | 5 | 2 | 0 | 2 | 1 | 2 | 3 | 3 | 80 |
| Total | 1 | 5 | 1 | 1 | 67 | 41 | 1 | 2 | 5 | 2 | 1 | 2 | 1 | 16 | 3 | 3 | 152 |

**Table 2:** Country of exposure of the 152 SARS-CoV-2 genomes with patient status information.

| Severity \ Country | Austria | Brazil | China | Colombia | Egypt | Gambia | Hong Kong | India | Indonesia | Israel | Italy | Japan | Malaysia | Mexico | Sri Lanka | UK | USA | Total |
| --- | --- | --- | --- | --- | --- | --- | --- | --- | --- | --- | --- | --- | --- | --- | --- | --- | --- | --- |
| Asymptomatic |  | 1 | 1 |  |  |  |  | 6 |  |  | 1 | 60 |  |  |  | 1 | 2 | 72 |
| Symptomatic | 16 | 1 | 1 | 6 | 10 | 1 | 3 | 4 | 1 | 3 |  | 17 | 1 | 1 | 1 |  | 14 | 80 |
| Total | 16 | 2 | 2 | 6 | 10 | 1 | 3 | 10 | 1 | 3 | 1 | 77 | 1 | 1 | 1 | 1 | 16 | 152 |

### Genetic variations at nucleotide position 11,083 is associated with COVID-19 severity

To identify potential viral genetic variations associated with the COVID-19 severity, we performed a GWAS by using TreeWAS [24] – a phylogenetic-based approach for GWAS of microbial genomes, which can take into account the observed virus population structure. The estimated ML tree (**Figure 1**) was used for the population structure correction. Our analysis identified genetic variations at the nucleotide position 11,083 (with respect to the reference SARS-CoV-2 genome sequence, accession number: NC_045512) to be significantly associated with the disease severity (**Figure 2**). By applying this analysis to all bootstrap trees, we found that this genomic position was identified 751/1,000 = 75.1% of the times, suggesting that our result was robust to the population structure uncertainty.

**Figure 2.**
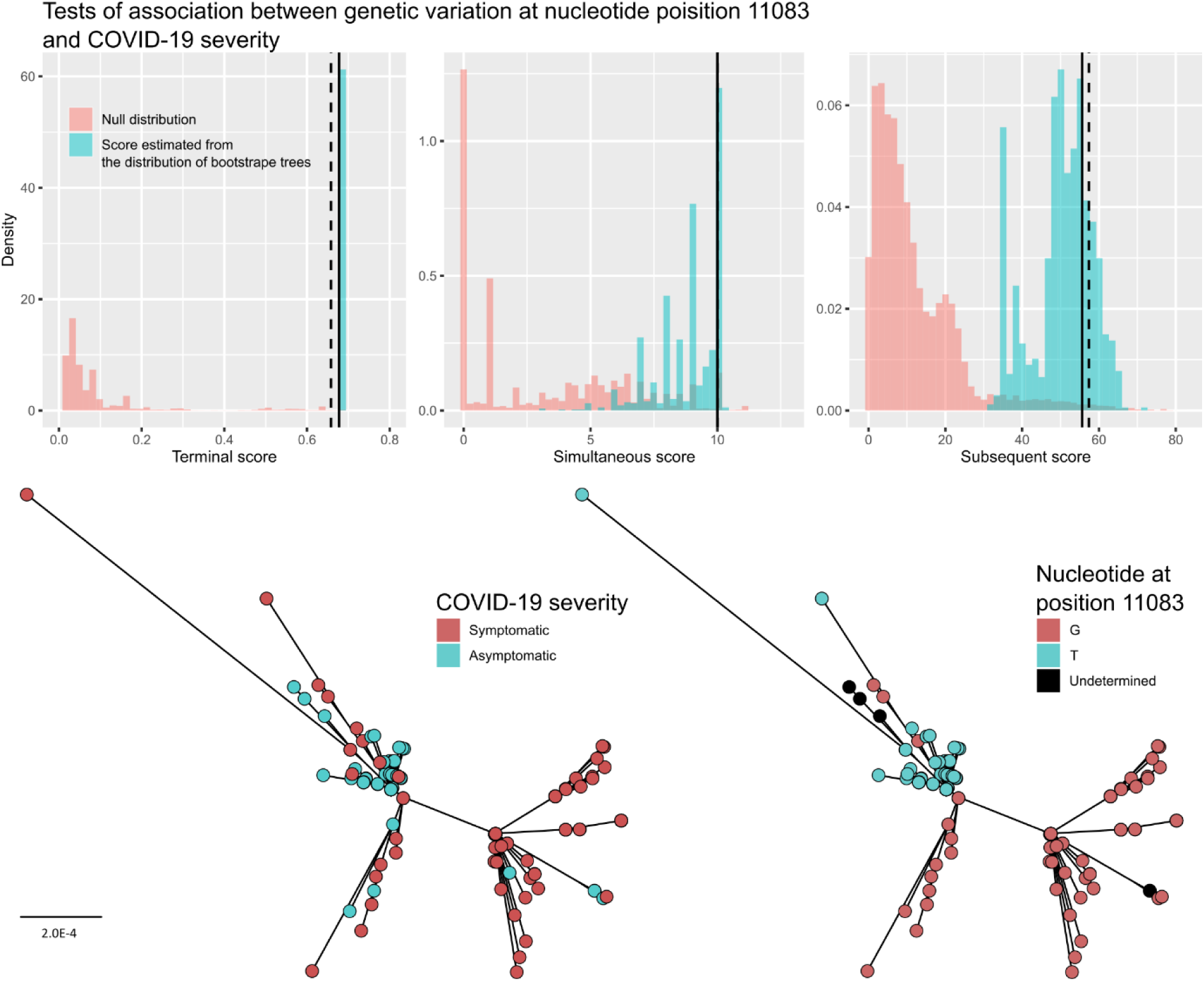
Genetic variations at nucleotide position 11,083 is associated with COVID-19 severity. (**Top**) three tests of genome-wide association analysis were performed, namely “Terminal test” (left), “Simultaneous test”, and “Subsequent test” to identify potential genetic variations associated with COVID-19 severity by using TreeWAS [24]. Across all three analyses, we identified genetic variations at the nucleotide position 11,083 (with respect to the reference SARS-CoV-2 genome, NC_045512) to be associated with COVID-19 severity. The null distributions of the scores are shown in red. The scores estimated based on the consensus tree (**Figure 1**) are indicated by black vertical lines. The significant thresholds corrected for multiple testing are indicated by dotted black vertical lines. The distributions of the scores estimated from the 1,000 bootstrap trees are shown in blue. Trees were pruned to contain only those with patient status before the analysis as recommended. (**Bottom**) the maximum likelihood tree of SARS-CoV-2 pruned to contain only those with patient status (152 sequences). The scales bar is in the units of substitutions per site. The tips were coloured by COVID-19 severity (**left**) or by the nucleotide variant at the position 11,083 (**right**).

Two genetic variations were observed at the nucleotide position 11,083, namely thymine (11083T, 75/152 = 49.34%) and guanine (11083G, 72/152 = 47.37%). 5 sequences (5/152 = 3.29%) had undetermined nucleotides at this site. We found that asymptomatic SARS-CoV-2 tended to have 11083T (N(11083T)/N(11083G) = 60/7), while viruses causing symptomatic cases tended to have 11083G (N(11083T)/N(11083G) = 15/65) (**Figure 2**). The relative risk ratio of developing symptoms given 11083G to 11083T is (65/72)/(15/75) = 4.51 times (95% confident interval = 2.85–7.14), and the odd ratio was estimated to be 37.14 by the Wald method (95% confident interval = 14.17–97.33). A study of SARS-CoV-2 from a Shanghai cohort has indeed identified the 11083T variant to be more prevalent in asymptomatic cases (9 in 91 cases = 9.89%) compared to symptomatic cases (1 in 21 cases = 4.76%), but the association was not significant [19]. This was likely due to their relatively small data set (N = 112), and different groupings of the disease outcome (where mild symptomatic and asymptomatic cases were grouped together as one and was compared against severe and critical cases), which could potentially mask the effect we observed. Another study identified this nucleotide position as being under a positive selection pressure [25], consistent with our finding. The closest relative of SAR-CoV-2 currently known is the bat coronavirus RaTG13 (GenBank accession number: MN996532) [26], and it has a G at this position, suggesting that G is the ancestral state. We hence designated this mutation 11083G>T. This mutation locates in the coding region of non-structural protein 6 (nsp6, nt 10973–11842), coded by ORF1ab (nt 266–21,555). 11083G>T is a non-synonymous mutation, conferring an amino acid change from leucine (L) to phenylalanine (F) in the nsp6 protein (L37F).

### 11083G and 11083T variants may interact with human miRNAs differently

Interaction between host microRNAs (miRNAs) and viral transcripts has been implicated in pathogenesis of viral diseases [27]. In this study, we examined whether the two virus variants have different modes of binding to human miRNAs by querying their nucleotide sequences against the miRNA database miRDB [28]. Although a coronavirus genome is a positive-sense RNA, negative-sense genomic RNAs are nonetheless generated during their replication, presenting a possibility that it might also interact with human miRNAs. We therefore examined both positive- and negative-strands of SARS-CoV-2 genome sequences.

On the positive strand, two miRNAs were predicted to uniquely target the 11083G with the same (highest) scores, namely miR-485-3p (5’–GUCAUACACGGCUCUCCUCUCU-3’) and miR-539-3p (5’–AUCAUACAAGGACAAUUUCUUU-3’). These two miRNAs belong to the same miRNA cluster (chr14: 101,047,321–101,047,398, and chr14: 101,055,419–101,055,491, respectively), and share identical seed sequences (5’-UCAUACA-3’), spanning the nt 11,083 (11,082–11,088) (**Figure 3**). By using the negative-sense sequence as a query, miR-3149 (5’–UUUGUAUGGAUAUGUGUGUGUAU-3’) was predicted to bind to the 11083G variant, and not the 11083T variant (nt c11087–11080). Our analyses did not identify miRNAs that specifically target the 11083T variant but not the 11083G variant. This suggested that the two variants might interact with these three miRNAs differently.

**Figure 3.**
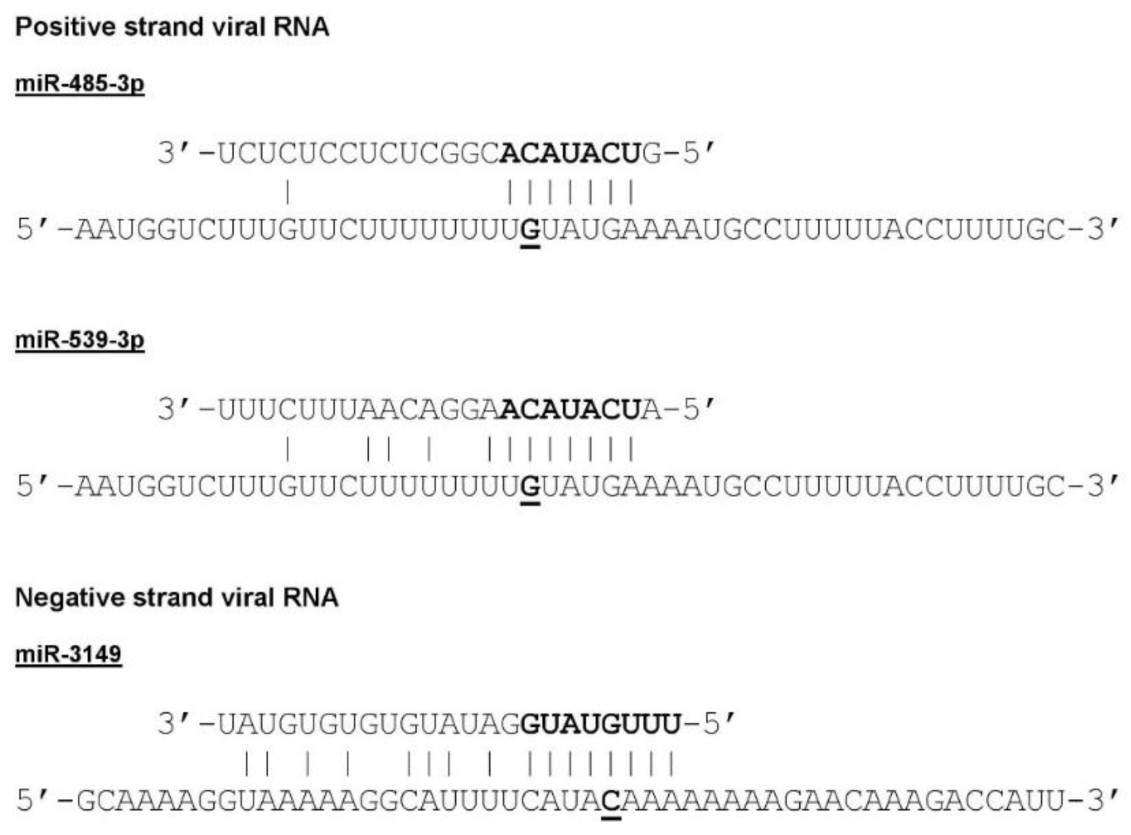
miR-485-3p, and miR-539-3p, and miR-3149 miRNAs specially target the 11083G variant of SARS-CoV-2, but not the 11083T variant. Sequences of the two SARS-CoV-2 variants as defined by the genetic variations at the nucleotide position 11,083 (11083G and 11083T variants) were searched against the miRNA database miRDB [28]. We found that miR-485-3p, and miR-539-3p were predicted to specially target the 11083G variant and not the 11083T variant on the positive strand. miR-3149 was predicted to uniquely target the 11083G variant on the negative strand. The viral genetic variations are written in bold and underlined. The seed sequences of miRNAs are shown in bold.

### Potential biological significance of 11083G>T mutation in COVID-19 severity

A study demonstrated that miR-485 can down regulate antiviral immunity by interacting with *retinoic acid–inducible gene I (RIG-I)* mRNA [29], the protein product of which can sense viral RNAs inside the cell, and activates the cell antiviral immune response. It is possible that the two variants of SARS-CoV-2 may interact with miR-485-3p differently, leading to differential host antiviral immune response, and subsequently differential COVID-19 severity. Moreover, one of the most common symptoms of severe COVID-19 is the cytokine release syndrome [30,31]. RIG-I signalling pathway is known to induce production of TNFα [32], which is a pro-inflammatory cytokine involved in the cytokine release syndrome [30]. It is thus also possible that the RNA molecules produced by the 11083G variant might sequester miR-485-3p, and ultimately lead to over-production of TNFα through an unregulated upregulation of the RIG-I pathway, resulting in a severe COVID-19 disease. This result warrants further experimental investigations how the SARS-CoV-2 11083G variant interacts with miR-485-3p.

Regarding miR-539-3p, a recent study showed that it suppresses expression of the pro-angiogenic factor Jagged1 [33] – a ligand for the Notch signalling pathway which controls cell proliferation and differentiation of various cell lineages, including blood vessel formation and sprouting [34,35]. Similar to the case of miR-485-3p, the viral RNAs produced by the 11083G variant might sequester miR-539-3p, leading to up-regulation of Jagged1 and subsequently angiogenesis, as observed in several symptomatic COVID-19 patients [36]. In addition to cell proliferation, miR-539 is also known to upregulate autophagy [37], the process by which cells degrade and recycle cellular components [38]. Incidentally, nsp6, the viral protein which the 11083G>T mutation directly affects, has also been reported to interfere with the host autophagy, restricting autophagosome expansion [39,40]. This restriction of autophagosome size likely compromises the cell ability to deliver viral components to lysosomes for degradation, and hence favouring virus infection [39]. Together, our results suggested potential roles of autophagy in pathogenesis of SARS-CoV-2, and should be further investigated.

In comparison to miR-485-3p and miR-539-3p, the functions of miR-3149 are much less known and well-understood. Nevertheless, it has been reported that patients with acute coronary syndrome had high levels of miR-3149 in their plasma [41]. Whether miR-3149 plays a role in COVID-19 pathogenesis is yet to be determined. Similarly, it is still unclear how the 11083T variant interacts with the host and causes an asymptomatic infection. Functional investigation of this mutation is warranted.

## Conclusion and final remarks

An unprecedented number of SARS-CoV-2 genomes have been generated at a rapid rate and made publicly available in near real-time like never before. To date, there are more than 30,000 sequences of SARS-CoV-2 genomes made publicly available on the GISAID database [20], and many of these sequences have patient information available. This allowed us to investigate viral genetic factors that might be associated with the COVID-19 severity.

In this study, we performed a GWAS on 152 SARS-CoV-2 genomes, and identified nucleotide variations at the genomic position 11,083 to be associated with the disease severity. Most of symptomatic cases were found to be infected with the 11083G variant, while the 11083T variant appeared to be associated more often with asymptomatic infections (relative risk ratio = 4.51 (2.85–7.14); odd ratio = 37.14 (14.17–97.33)). The two nucleotide variants, 11083G and 11083T, are non-synonymous, corresponding to L and F at the amino acid position 37 in the nsp6 protein, respectively. Our results have potential applications for the development of better, and more informative test kits, potentially allowing for asymptomatic cases to be distinguished from symptomatic cases. Continual surveillance of COVID-19 should monitor this genomic region as well as its surrounding neighbourhood as they might affect the pathogenesis of COVID-19.

Bioinformatic analyses suggested that the two variants might interact with the human host miRNAs differently. In particular, the 11083G variant was identified as a potential target of miR-485-3p, miR-539-3p, and miR-3149, while the 11083T variant was not. These differences might contribute to the observed differential association between the two variants and the disease severity. Our results warrant further experimental confirmations to validate biological significance of these genetic variations and their consequences.

## Methods

### SARS-CoV-2 genome sequences with patient status

Genome sequences of SARS-CoV-2 with patient status were downloaded from the GISAID database [20] on 18/05/2020 with their metadata. To allow for accurate determination of genetic factors associated with COVID-19 severity, we only analysed sequences whose patient status could be unambiguously determined as either “asymptomatic” or “symptomatic”. We designated a virus to cause an asymptomatic infection if its patient status was either “Asymptomatic/Released” or “asymptomatic”. A virus was determined to cause a symptomatic infection if the patient status was either “Hospitalized in ICU”, “Hospitalized/Deceased”, “ICU; Serious”, “Intensive Care Unit”, “pneumonia (chest X-ray)”, “Severe/ICU”, or “Symptomatic”. Sequences that were not generated from original clinical samples were excluded. Those with ambiguous nucleotides greater than 5% of the total sequence length, and whose total lengths were less than 29,500 nucleotides were also excluded from downstream analyses. Although EPI_ISL_417919 was found to fit the inclusion criteria, manual inspection revealed that it contained many unique nucleotide variants surrounding its multiple undetermined regions of “N”s, likely due to sequencing and / or assembly errors, and had about 4.28% of undetermined nucleotides. It was thus also excluded from our dataset. In total, our dataset comprised 152 sequences. A table of acknowledgements for the sequences used in this study can be found in **Table S1**.

### Lineage assignment

The lineage of all genomes were determined based on the methodology described in [21] by using pangolin (github.com/hCoV-2019/pangolin) with the reference lineage version 07/05/2020 under the default setting. All prediction passed the quality control.

### Phylogenetic analysis

500 randomly sampled SARS-CoV-2 genomes were downloaded from GISAID on 21/05/2020 (**Table S1**), all of which were full-length and had a high sequencing coverage. Together with the 152 genomes with patient status, a manually-curated multiple sequence alignment of 652 SARS-CoV-2 genomes was constructed. Potential recombination within the alignment was checked by using RDP, GENECONV, Chimera, MaxChi, and 3Seq, all implemented in Recombination Detection Program 4 [42]. Sites with more than 50% ambiguous nucleotides were excluded from recombination analysis. Sites with the most common nucleotide present in more than 99% of the sequences were also excluded. None of the program found evidence for recombination events within the data, suggesting that our dataset was recombinant-free.

A maximum likelihood phylogeny was estimated from the prepared full-length alignment by using IQ-TREE [22]. The best-fit nucleotide substitution models was determined to be GTR^+^I^+^F under the Bayesian information criterion by using ModelFinder [23] and was used for the tree reconstruction. The bootstrap clade support was computed by using 1,000 pseudoreplicate datasets.

### Identification of genetic variations associated with COVID-19 severity

TreeWAS [24] was used to identify potential genetic variations associated with COVID-19 severity, defined as two discrete traits: “asymptomatic” and “symptomatic”. The estimated ML tree was used for the population structure correction. The tree was rooted by assuming that lineage A and B of SARS-CoV-2 are monophyletic, and was pruned to contain only those with patient status before the analysis as recommended. Three tests of association were performed, including the “terminal”, “simultaneous”, and “subsequent” tests. Sites with more than 50% ambiguous nucleotides were removed. Sites with the most common nucleotide present in more than 95% of the sequences were also removed. Only 15 variant loci remained after the filtering. Ancestral states of both genetic and phenotypic data were inferred under a maximum likelihood framework as implemented in the package. The number of sites simulated for estimating the null distribution was 1,000×15 = 15,000 sites. The overall threshold of significance was set to 5%, and corrected to be 5%/15/3 = 0.11% in each test under the Bonferonni multiple-testing correction criteria as recommended. An association was considered significant if it was detected by at least one of the three tests aforementioned. We also apply this analysis to all of the 1,000 trees in the bootstrap tree distribution obtained from the phylogenetic analysis described above to examine the robustness of the result. Our analyses robustly identified nucleotide variations at the genomic position 11,083 (with respect to the reference SARS-CoV-2 genome, NC_045512) to be significantly associated with the disease severity.

### microRNA analyses

To examine potential biological significance of the identified genetic variations, we searched the two sequence variants against the microRNA database miRDB [28] under the default settings, available at http://mirdb.org/.

## Data Availability

All sequence data used in this study were retrieved from GISAID. The table of acknowledgement of the sequences used can be found in Table S1.

## Data Availability

All sequence data used in this study were retrieved from GISAID. The table of acknowledgement of the sequences used can be found in **Table S1**.

## Acknowledgments

This research project is partially supported by Mahidol University (MRC-IM 02/2563).

## Author contributions

P.A., P.W., A.T. conceived the study. P.A., Y.T, P.W. performed data curation, analysis, and interpretation of the results. P.A., P.W. drafted the manuscript. All revised and approved of the final manuscript.

## Competing Interests

The authors declare no conflict of interest.

## Notes

### Competing Interest Statement

The authors have declared no competing interest.

### Funding Statement

This research project is supported by Mahidol University (MRC-IM 02/2563).

## References

1. Zhu, N.; Zhang, D.; Wang, W.; Li, X.; Yang, B.; Song, J.; Zhao, X.; Huang, B.; Shi, W.; Lu, R.; et al. A novel coronavirus from patients with pneumonia in China, 2019. N. Engl. J. Med. 2020.

2. International Society for Infectious Diseases PRO/AH/EDR> Undiagnosed pneumonia - China (HU): RFI 2019, 20191230.6864153.

3. Gorbalenya, A.E.; Baker, S.C.; Baric, R.S.; de Groot, R.J.; Drosten, C.; Gulyaeva, A.A.; Haagmans, B.L.; Lauber, C.; Leontovich, A.M.; Neuman, B.W.; et al. The species Severe acute respiratory syndrome-related coronavirus: Classifying 2019-nCoV and naming it SARS-CoV-2. Nat. Microbiol. 2020, 5, 536–544.

4. Huang, C.; Wang, Y.; Li, X.; Ren, L.; Zhao, J.; Hu, Y.; Zhang, L.; Fan, G.; Xu, J.; Gu, X. Clinical features of patients infected with 2019 novel coronavirus in Wuhan, China. Lancet 2020, 395, 3–506.

5. Chen, N.; Zhou, M.; Dong, X.; Qu, J.; Gong, F.; Han, Y.; Qiu, Y.; Wang, J.; Liu, Y.; Wei, Y.; et al. Epidemiological and clinical characteristics of 99 cases of 2019 novel coronavirus pneumonia in Wuhan, China: a descriptive study. Lancet 2020.

6. World Health Organization Statement on the second meeting of the International Health Regulations (2005) Emergency Committee regarding the outbreak of novel coronavirus (2019-nCoV) Available online: https://www.who.int/news-room/detail/30-01-2020-statement-on-the-second-meeting-of-the-international-health-regulations-(2005)-emergency-committee-regarding-the-outbreak-of-novel-coronavirus-(2019-ncov).

7. World Health Organization WHO Director-General’s opening remarks at the media briefing on COVID-19 - 11 March 2020 Available online: https://www.who.int/dg/speeches/detail/who-director-general-s-opening-remarks-at-the-media-briefing-on-covid-19-11-march-2020.

8. Worldometers.info COVID-19 CORONAVIRUS PANDEMIC Available online: https://www.worldometers.info/coronavirus/ (accessed on May 22, 2020).

9. Guan, W.J.; Ni, Z.Y.; Hu, Y.; Liang, W.H.; Ou, C.Q.; He, J.X.; Liu, L.; Shan, H.; Lei, C.L.; Hui, D.S.C.; et al. Clinical characteristics of coronavirus disease 2019 in China. N. Engl. J. Med. 2020, 382, 3–1720.

10. Chan-Yeung, M.; Xu, R.H. SARS: Epidemiology. Respirology 2003, 8, S9–S14.

11. World Health Organization Consensus document on the epidemiology of severe acute respiratory syndrome Available online: https://apps.who.int/iris/handle/10665/70863.

12. World Health Organization Middle East respiratory syndrome coronavirus (MERS-CoV) Available online: https://www.who.int/emergencies/mers-cov/en/.

13. Lauer, S.A.; Grantz, K.H.; Bi, Q.; Jones, F.K.; Zheng, Q.; Meredith, H.R.; Azman, A.S.; Reich, N.G.; Lessler, J. The incubation period of coronavirus disease 2019 (COVID-19) from publicly reported confirmed cases: Estimation and application. Ann. Intern. Med. 2020, 172, 3–582.

14. Heneghan, C.; Brassey, J.; Jefferson, T. COVID-19: What proportion are asymptomatic? Available online: https://www.cebm.net/covid-19/covid-19-what-proportion-are-asymptomatic/.

15. Furukawa, N.W.; Brooks, J.T.; Sobel, J. Evidence supporting transmission of severe acute respiratory syndrome coronavirus 2 while presymptomatic or asymptomatic. Emerg. Infect. Dis. 2020.

16. Yu, X.; Yang, R. COVID-19 transmission through asymptomatic carriers is a challenge to containment. Influenza Other Respi. Viruses 2020.

17. Gandhi, M.; Yokoe, D.S.; Havlir, D. V. Asymptomatic transmission, the Achilles’ heel of current strategies to control covid-19. N. Engl. J. Med. 2020.

18. Yang, X.; Yu, Y.; Xu, J.; Shu, H.; Xia, J.; Liu, H.; Wu, Y.; Zhang, L.; Yu, Z.; Fang, M.; et al. Clinical course and outcomes of critically ill patients with SARS-CoV-2 pneumonia in Wuhan, China: a single-centered, retrospective, observational study. Lancet Respir. Med. 2020, 8, 3–481.

19. Zhang, X.; Tan, Y.; Ling, Y.; Lu, G.; Liu, F.; Yi, Z.; Jia, X.; Wu, M.; Shi, B.; Xu, S.; et al. Viral and host factors related to the clinical outcome of COVID-19. Nature 2020.

20. Shu, Y.; McCauley, J. GISAID: Global initiative on sharing all influenza data - from vision to reality. Eurosurveillance 2017, 22, 30494.

21. Rambaut, A.; Holmes, E.C.; Hill, V.; OToole, A.; McCrone, J.; Ruis, C.; Plessis, L. du; Pybus, O. A dynamic nomenclature proposal for SARS-CoV-2 to assist genomic epidemiology. bioRxiv 2020, 2020.04.17.046086.

22. Nguyen, L.T.; Schmidt, H.A.; Von Haeseler, A.; Minh, B.Q. IQ-TREE: A fast and effective stochastic algorithm for estimating maximum-likelihood phylogenies. Mol. Biol. Evol. 2015, 32, 3–274.

23. Kalyaanamoorthy, S.; Minh, B.Q.; Wong, T.K.F.; Von Haeseler, A.; Jermiin, L.S. ModelFinder: Fast model selection for accurate phylogenetic estimates. Nat. Methods 2017, 14, 3–589.

24. Collins, C.; Didelot, X. A phylogenetic method to perform genome-wide association studies in microbes that accounts for population structure and recombination. PLoS Comput. Biol. 2018, 14, e1005958.

25. Benvenuto, D.; Angeletti, S.; Giovanetti, M.; Bianchi, M.; Pascarella, S.; Cauda, R.; Ciccozzi, M.; Cassone, A. Evolutionary analysis of SARS-CoV-2: How mutation of Non-Structural Protein 6 (NSP6) could affect viral autophagy. J. Infect. 2020.

26. Zhou, P.; Yang, X. Lou; Wang, X.G.; Hu, B.; Zhang, L.; Zhang, W.; Si, H.R.; Zhu, Y.; Li, B.; Huang, C.L.; et al. A pneumonia outbreak associated with a new coronavirus of probable bat origin. Nature 2020, 579, 3–273.

27. Bruscella, P.; Bottini, S.; Baudesson, C.; Pawlotsky, J.M.; Feray, C.; Trabucchi, M. Viruses and miRNAs: More friends than foes. Front. Microbiol. 2017, 8, 824.

28. Chen, Y.; Wang, X. miRDB: an online database for prediction of functional microRNA targets. Nucleic Acids Res. 2020, 48, D127-D131.

29. Ingle, H.; Kumar, S.; Raut, A.A.; Mishra, A.; Kulkarni, D.D.; Kameyama, T.; Takaoka, A.; Akira, S.; Kumar, H. The microRNA miR-485 targets host and influenza virus transcripts to regulate antiviral immunity and restrict viral replication. Sci. Signal. 2015, 8, ra126.

30. Moore, B.J.B.; June, C.H. Cytokine release syndrome in severe COVID-19. Science (80-.). 2020, 368, 3–474.

31. Huang, C.; Wang, Y.; Li, X.; Ren, L.; Zhao, J.; Hu, Y.; Zhang, L.; Fan, G.; Xu, J.; Gu, X.; et al. Clinical features of patients infected with 2019 novel coronavirus in Wuhan, China. Lancet 2020, 395, 3–506.

32. Wang, J.; Wu, S.; Jin, X.; Li, M.; Chen, S.; Teeling, J.L.; Perry, V.H.; Gu, J. Retinoic acid-inducible gene-I mediates late phase induction of TNF-α by lipopolysaccharide. J. Immunol. 2008, 180, 3–8019.

33. Su, H.; Wang, X.; Song, J.; Wang, Y.; Zhao, Y.; Meng, J. MicroRNA-539 inhibits the progression of Wilms’ Tumor through downregulation of JAG1 and Notch1/3. Cancer Biomarkers 2019, 24, 3–133.

34. Kume, T. Novel insights into the differential functions of Notch ligands in vascular formation. J. Angiogenes. Res. 2009, 1, 8.

35. Lin, J.; Lin, Y.; Su, L.; Su, Q.; Guo, W.; Huang, X.; Wang, C.; Lin, L. The role of Jagged1/Notch pathway-mediated angiogenesis of hepatocarcinoma cells in vitro, and the effects of the spleen-invigorating and blood stasis-removing recipe. Oncol. Lett. 2017, 14, 3–3622.

36. Ackermann, M.; Verleden, S.E.; Kuehnel, M.; Haverich, A.; Welte, T.; Laenger, F.; Vanstapel, A.; Werlein, C.; Stark, H.; Tzankov, A.; et al. Pulmonary vascular endothelialitis, thrombosis, and angiogenesis in Covid-19. N. Engl. J. Med. 2020.

37. Hui, J.; Huishan, W.; Tao, L.; Zhonglu, Y.; Renteng, Z.; Hongguang, H. miR-539 as a key negative regulator of the MEK pathway in myocardial infarction. Herz 2017, 42, 3–789.

38. Mizushima, N.; Komatsu, M. Autophagy: Renovation of cells and tissues. Cell 2011, 147, 3–741.

39. Cottam, E.M.; Whelband, M.C.; Wileman, T. Coronavirus NSP6 restricts autophagosome expansion. Autophagy 2014, 10, 3–1441.

40. Cottam, E.M.; Maier, H.J.; Manifava, M.; Vaux, L.C.; Chandra-Schoenfelder, P.; Gerner, W.; Britton, P.; Ktistakis, N.T.; Wileman, T. Coronavirus nsp6 proteins generate autophagosomes from the endoplasmic reticulum via an omegasome intermediate. Autophagy 2011, 7, 3–1347.

41. Li, X.; Yang, Y.; Wang, L.; Qiao, S.; Lu, X.; Wu, Y.; Xu, B.; Li, H.; Gu, D. Plasma miR-122 and miR-3149 potentially novel biomarkers for acute coronary syndrome. PLoS One 2015, 10, e0125430.

42. Martin, D.P.; Murrell, B.; Golden, M.; Khoosal, A.; Muhire, B. RDP4: Detection and analysis of recombination patterns in virus genomes. Virus Evol. 2015, 1, vev003.

